# Comparable Efficacy of Curcumin and Proton Pump Inhibitor for Functional Dyspepsia: A Randomized Double-blind Controlled Trial

**DOI:** 10.1101/2022.12.08.22283167

**Authors:** Pradermchai Kongkam, Wichittra Khongkha, Chawin Lopimpisuth, Chisanucha Chumsri, Prach Kosarussawadee, Phanupong Phutrakool, Sittichai Khamsai, Kittisak Sawanyawisuth, Thanyachai Sura, Pochamana Phisalprapa, Thanwa Buamahakul, Sarawut Siwamogsatham, Jaenjira Angsusing, Pratchayanan Poonniam, Kulthanit Wanaratna, Monthaka Teerachaisakul, Krit Pongpirul

## Abstract

**Objectives:** Curcumin has been claimed to have gastrointestinal benefits, including dyspepsia, a common disorder that could be managed in a primary care setting with behavioral and dietary modifications as well as over-the-counter medications. This study aimed to compare the efficacy of curcumin versus omeprazole in improving patient-reported outcomes.

**Design:** Patients with functional dyspepsia were randomized to curcumin alone (C), omeprazole alone (O), or curcumin plus omeprazole (C+O). Patients in the combination group received 2 capsules of 250 mg curcumin 4 times daily and 1 capsule of 20 mg omeprazole once daily for 28 days. The primary outcomes were functional dyspepsia symptoms on days 28 and 56 assessed using the Severity of Dyspepsia Assessment (SODA) scores. Secondary outcomes were the occurrence of adverse events (AEs) and serious adverse events (SAEs).

**Results:** A total of 206 enrolled patients were randomly assigned to the three groups, of which 151 completed the study. Demographic data (age 49.7±11.9 years; female 73.4%), clinical characteristics, and baseline dyspepsia scores were comparable between the three groups. SODA scores in each group showed significant improvement on day 28 and day 56 in the pain, non-pain, and satisfaction categories. No significant differences were observed among the three groups and no serious adverse events occurred.

**Conclusion:** Curcumin and omeprazole have comparable efficacy for functional dyspepsia with no obvious synergistic effect.

**Key Messages:** *What is already known:*

- Dyspepsia is one of the most common disorders in which patients usually try behavioral and diet modifications, and over-the-counter drugs before seeing a physician.
- Proton pump inhibitors have been established as an effective treatment for functional dyspepsia.
- Curcumin, an active ingredient in turmeric, is currently used for the treatment of dyspepsia in countries in Southeast Asia.
- However, there is no clinical trial evidence to support the use of curcumin as a first-line treatment.

*What this study adds:*

- Based on this double-blind, placebo-controlled clinical trial, oral curcumin is safe and well tolerated.
- Patients with functional dyspepsia were treated with curcumin, curcumin plus omeprazole, and omeprazole, which showed a similar significant symptomatic improvement.
- There was no synergistic effect detected between omeprazole and curcumin.

*How this study might affect research, practice, or policy:*

- This study provides a possible additional drug option for patients with functional dyspepsia.
- More clinical trials are required to assess long-term outcomes and adverse events.

## INTRODUCTION

Dyspepsia is a frequently occurring disorder that can be caused by a variety of factors, with no evidence of other structural diseases that exhibit similar symptoms. Although dyspepsia is common, most patients do not schedule an appointment with a doctor to treat this condition. One-quarter of patients with dyspepsia have symptoms that require specific treatment, while the rest do not have the symptoms that define them as functional dyspepsia (1). According to Rome IV criteria, patients diagnosed with functional dyspepsia have postprandial fullness, early satiation, epigastric pain or burning, and no evidence of structural disease (including at upper endoscopy) to explain the symptoms. Postprandial Distress Syndrome (PDS) and Epigastric Pain Syndrome (EPS) are two types of functional dyspepsia (2).

Most patients with functional dyspepsia in a primary care setting would be treated with behavioral and dietary modifications, as well as over-the-counter medications, before seeing a physician. Over-the-counter proton pump inhibitors (PPIs) are commonly recommended in several countries, whereas patients with persistent symptoms require medical attention for possible Helicobacter pylori infection (3-5). The recent Cochrane systematic review on the use of PPIs for functional dyspepsia showed better overall effectiveness with the number needed to treat 11 (6). However, prolonged use of PPIs demonstrated a potential increased risk of fractures, micronutrient deficiencies, infection, etc. Due to the lack of a higher-quality study, these possible adverse effects remain controversial (7-9).

Turmeric, scientifically named *Curcuma longa* L., is a plant that has been used extensively for a long period. The active ingredient of its rhizome, curcumin, is used medically topically and orally. In addition to nourishing creams and cosmetics, curcumin powder capsules have been used for the treatment of dyspepsia and other gastrointestinal problems. Turmeric is one of the herbs that is frequently used to alleviate symptoms similar to dyspepsia among Thai people and those who live near Thailand. However, few conventional physicians have chosen this herbal medicine as the first drug of choice against functional dyspepsia. This is in part due to a lack of research comparing the efficacy and side effects of curcumin with PPI in the treatment of functional dyspepsia. This study aimed to compare the efficacy of curcumin versus PPI in the treatment of patients with functional dyspepsia in a placebo-controlled double-blind trial.

## METHODS

### Study Design

This multicenter, randomized, double-blind, placebo-controlled, parallel-group trial was conducted at the Thai Traditional Medicine Institute and Chao Phraya Abhaibhubejhr Hospital from June 2019 to April 2020. The patients were randomly assigned into three groups: curcumin+omeprazole (C+O), curcumin only (C only), and omeprazole only (O only).

### Study Population

Patients who were willing to participate in this trial were evaluated for eligibility: symptoms compatible with functional dyspepsia, between 18 and 70 years, ECOG performance status 0 or 1, not taken aspirin or NSAIDs for the past 3 months, not taken curcumin or food that significantly contained curcumin during 4 weeks before the study, no symptoms of irritable bowel syndrome (constipation, diarrhea, and frequent defecation), not taken herbal medication or medication that can affect gastrointestinal symptoms and diseases, and not taking PPIs during the 4 weeks before the study. Patients who were pregnant/breastfeeding; had a curcumin allergy; had gallstones; had ulcers or lumps in their stomach, duodenum, or esophagus; had severe inflammation of the gastric mucosa oesophageal mucosa or intestinal mucosa; had been infected with gastric *Helicobacter pylori*; had diseases that would hinder the treatment of functional dyspepsia, or had symptoms or physical signs that are a warning sign of serious diseases incompatible with functional dyspepsia, were excluded from this study. All patients underwent gastroscopy to confirm the functional dyspepsia diagnosis.

### Randomization and Blinding

Participants were recruited at the Institute of Thai Traditional Medicine, Department of Thai Traditional and Alternative Medicine, Ministry of Public Health, Bangkok, Thailand, and Chao Phraya Abhaibhubejhr Hospital, a tertiary care hospital in Prachinburi, Thailand. The eligible participants were randomly allocated into the three groups using a block randomization of size six at a ratio of 1:1:1. The identification numbers were generated and inserted into the opaque concealed envelope by researchers from the Institute of Thai Traditional Medicine, who were not involved in the care of the volunteers. Clinicians, data collectors, and patients were blinded during this entire process. The randomization identification number was assigned to three groups with the code of each participant and delivered sealed envelopes to the doctor conducting the research in the area.

### Recruitment

Methods for recruiting participants include being a patient under the care of physicians in both institutions, creating electronic media, printed media, promoting flyers, contacting the patient’s physician to introduce the researcher to the participants, and publicizing the study in healthcare facilities that provide both conventional and traditional Thai medicine services.

### Treatment and safety protocol

Each group of enrolled participants was provided with the medications in-person at both sites. The provided medications were packaged in two sizes: large capsules (250 mg of curcumin or placebo), and small capsules (20 mg of omeprazole or placebo). Each participant was instructed to take two large capsules four times a day and one small capsule once a day for 28 days.

Any possible side effects could be reported at any time, while patients were formally assessed for possible adverse events on day 28. The following discontinuation criterion was applied: (1) participants were allergic to the medication or unable to take the medication, (2) participants did not follow up on the evaluation of treatment, (3) participants were unable to tolerate the side effects of the medication, (4) pathological examination results of lesions in the esophagus, stomach, or duodenum, show compatibility with certain cancers or tumors, and (5) the endoscopy was unsuccessful. The study termination criteria included (1) serious adverse events (SAEs) of up to 3 percent or occurring in 6 or more volunteers and the matter has been brought to the ethics committee for consideration of the need to discontinue research and (2) doubts or serious ethical errors in the research process.

### Outcome Measurements

The Short-Form Leeds Dyspepsia Questionnaire (SF-LDQ) and the Severity of Dyspepsia Assessment (SODA) were used to measure the extent of the symptoms at the beginning of the study. Later, only SODA was used to observe the changes in severity on day 28 and day 56. Each measurement was performed in the clinics, except for that of day 56 which was conducted by telephone interview.

The secondary goal was to evaluate possible adverse events. On day 28, after evaluation, the patients were thoroughly examined by physical examination and interviews to determine whether an undesirable episode occurred during treatment.

### Sample Size Calculation

The sample size was calculated using a formula to compare the difference in scores by referring to Rosner and Bernard. However, the number of patients per group was 63, due to the possibility of loss, and in the case that participants have another disorder detected after endoscopy, which accounts for about 10%, the authors have therefore specified the sample size to be 70 people in each group.

### Ethics Consideration

This study was approved by the Ethics Committee for Research in Human Subjects in the Fields of Thai Traditional and Alternative Medicine (TAMEC No. 11-2561). Participants provided their written consent to participate in the study.

### Statistical Analysis

Descriptive and analytical statistics were performed by using STATA/MP statistical software release 15 (StataCorp LLC, College Station, TX). The generalized estimating equation (GEE) was used to analyze the primary outcomes at a statistical significance level of 0.05.

## RESULTS

A total of 241 patients were evaluated for eligibility; of these, 206 met the inclusion criteria and were enrolled and randomized into the three groups. The most common reason for ineligibility was incompatibility with the diagnosis of functional dyspepsia, followed by incorrect age, recent intake of PPI, being pregnant or breastfeeding, current *Helicobacter pylori* infection, and rejection of consent.

Overall, 69, 69, and 68 patients were randomly assigned to the C+O, C, and O groups; of these, 16, 20, and 19 dropped out, respectively. Detailed numbers and explanations for exclusion and dropout are shown in the CONSORT diagram (Figure 1). Baseline demographic and clinical characteristics were similar in the C+O, C, and O groups, of which the summation is given in Table 1.

**Table 1.** Characteristics of the Participants.

|  | C+O<br>(n=69) | C only<br>(n=69) | O only<br>(n=68) | p-value |
| --- | --- | --- | --- | --- |
| **Demographics** |  |  |  |  |
| - Female, n (%) | 49 (71.01) | 50 (72.46) | 52 (76.47) | 0.756 |
| - Age (years) | 49.54 ± 12.10 | 49.97 ± 11.96 | 49.3 ± 11.65 | 0.946 |
| - Height (cm) | 159.03 ± 8.62 | 158.8 ± 9.01 | 159.04 ± 7.56 | 0.982 |
| - Weight (kg) | 62.13 ± 12.19 | 62.71 ± 13.38 | 63.43 ± 12.36 | 0.835 |
| - BMI (kg/m<sup>2</sup>) | 24.44 ± 3.56 | 24.79 ± 4.43 | 25.05 ± 4.39 | 0.693 |
| **Vital signs** |  |  |  |  |
| - Pulse rate (beats/min) | 78.19 ± 12.01 | 78.38 ± 8.41 | 80.82 ± 10.24 | 0.253 |
| - Respiratory rate (beats/min) | 19.22 ± 1.46 | 20.03 ± 7.51 | 19.12 ± 1.40 | 0.433 |
| - Body temperature (°C) | 36.82 ± 0.37 | 36.81 ± 0.27 | 36.85 ± 0.26 | 0.705 |
| - Systolic blood pressure | 128.36 ± 13.85 | 129.96 ± 14.37 | 126.15 ± 16.13 | 0.321 |
| - Diastolic blood pressure | 77.65 ± 10.96 | 78.58 ± 9.69 | 75.96 ± 10.48 | 0.328 |
| **Dyspepsia group, n(%)** |  |  |  |  |
| - Postprandial Distress Syndrome (PDS) | 43 (63.24) | 42 (60.87) | 46 (67.65) | 0.704 |
| - Epigastric Pain Syndrome (EPS) | 25 (36.76) | 27 (39.13) | 22 (32.35) |  |
| **Laboratory** |  |  |  |  |
| - White blood cells (cells/mm<sup>3</sup>) | 6914.48 ± 2107.4 | 7401.08 ± 2199.63 | 7710.48 ± 2540.89 | 0.137 |
| - Hemoglobin (g/dL) | 13.56 ± 2.87 | 12.9 ± 1.40 | 12.42 ± 1.87 | 0.011 |
| - Hematocrit (%) | 40.04 ± 5.15 | 39.24 ± 3.16 | 39.12 ± 4.46 | 0.418 |
| - Platelet count (cells/mm<sup>3</sup>) | 260000 ± 57830.24 | 250000 ± 55400.04 | 270000 ± 93237.94 | 0.494 |
| - AST (SGOT) (U/L) | 26.76 ± 9.81 | 27.90 ± 13.38 | 25.43 ± 7.89 | 0.436 |
| - ALT (SGPT) (U/L) | 25.48 ± 16.96 | 25.64 ± 16.19 | 28.22 ± 22.11 | 0.658 |
| - Total bilirubin (mg/dL) | 1.11 ± 2.89 | 1.07 ± 2.91 | 1.04 ± 3.45 | 0.991 |
| - INR | 0.95 ± 0.15 | 0.99 ± 0.12 | 0.99 ± 0.1 | 0.113 |
| - Creatinine (mg/dL) | 0.81 ± 0.38 | 0.89 ± 0.59 | 0.91 ± 1.05 | 0.715 |
| - Sodium (mmol/L) | 139.5 ± 2.1 | 139.81 ± 2.38 | 137.59 ± 16.22 | 0.367 |
| - Potassium (mmol/L) | 3.96 ± 0.43 | 3.81 ± 0.43 | 3.92 ± 0.35 | 0.127 |
| - Chloride (mmol/L) | 104.08 ± 4.37 | 103.77 ± 3.9 | 103.5 ± 3.38 | 0.703 |
| - Total CO<sub>2</sub> (mmol/L) | 24.76 ± 2.93 | 25.61 ± 6.98 | 25.08 ± 3.32 | 0.598 |

**Figure 1.**
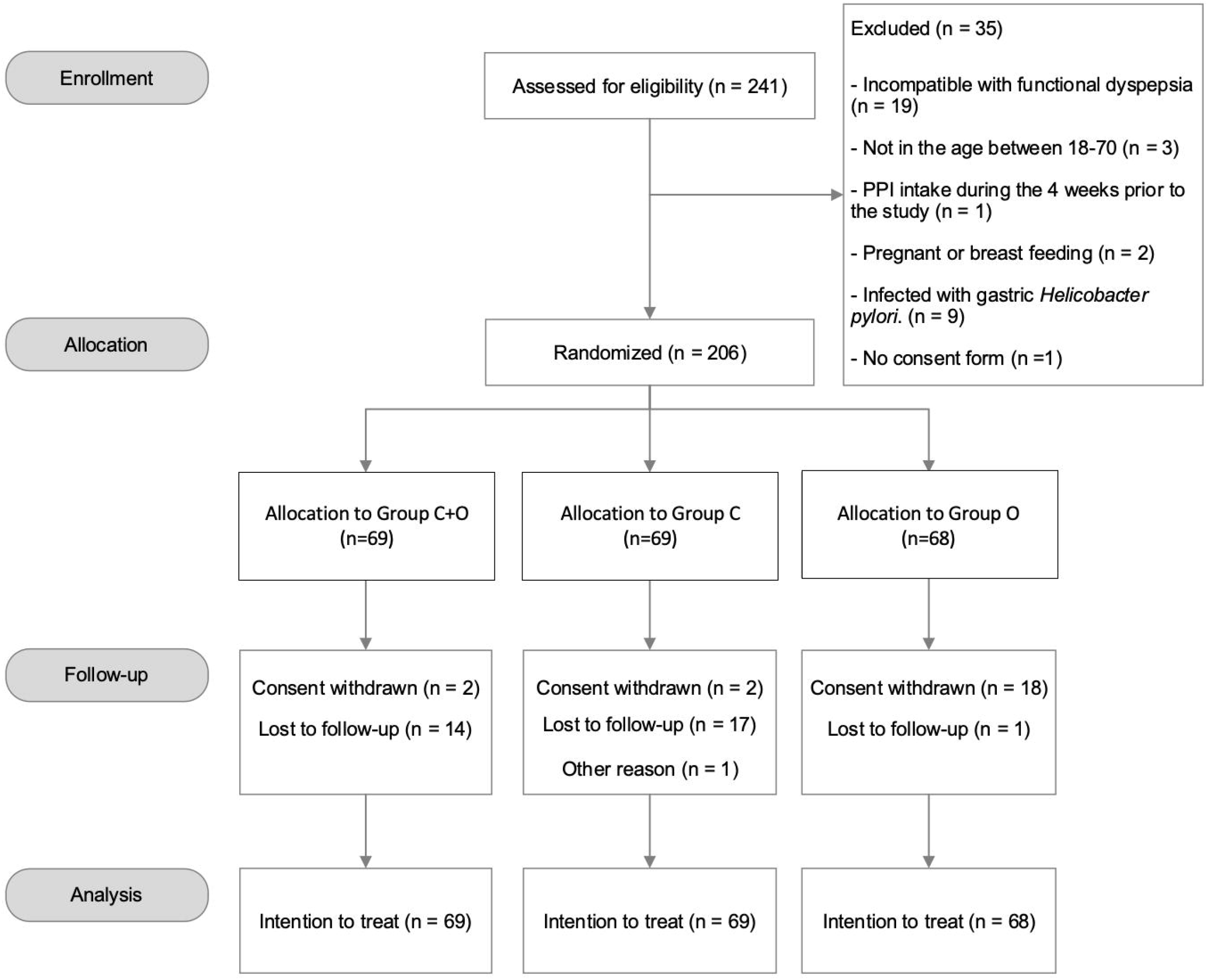
CONSORT Diagram.

### Dyspepsia symptom severity

At baseline, SF-LDQ did not show significant differences, whereas the overall SODA scores (pain intensity, non-pain symptoms, and satisfaction) were comparable between the three groups (Table 2). On day 28, a significant improvement in SODA pain intensity and non-pain symptoms scores was observed in three groups: the pain intensity decreased by −5.41 (95%CI −7.37, −3.45), −6.22 (95%CI −8.20, −4.25), and −6.98 (95%CI −8.95, −5.02) among the groups C+O, C and O, respectively, while the non-pain symptoms decreased by −2.41 (95%CI −3.29, −1.52), −2.45 (95%CI −3.34, −156), and −2.55 (95%CI −3.44, −1.66) among the groups C+O, C and O, respectively. On the other hand, the SODA satisfaction scores significantly improved by 0.86 (95%CI 0.01, 1.72) only in group C.

**Table 2.** Baseline SF-LDQ and SODA Scores.

| Factor | C+O<br>(n=69) | C only<br>(n=69) | O only<br>(n=68) | p-value |
| --- | --- | --- | --- | --- |
| **SODA** |  |  |  |  |
| - Pain intensity (2-47) | 25.80 ± 3.22 | 25.14 ± 4.09 | 26.26 ± 3.74 | 0.208 |
| - Non-pain symptoms (7-35) | 16.14 ± 3.18 | 15.58 ± 3.21 | 15.74 ± 2.69 | 0.530 |
| - Satisfaction (2-23) | 11.78 ± 1.98 | 11.84 ± 1.65 | 11.68 ± 2.29 | 0.891 |
| **SF-LDQ** |  |  |  |  |
| - Method 1 (Sum frequency) | 7.90 ± 4.23 | 7.01 ± 4.06 | 7.12 ± 3.56 | 0.360 |
| - Method 2 (Sum severity) | 7.97 ± 4.25 | 6.88 ± 4.14 | 6.85 ± 3.65 | 0.182 |
| - Method 3 (Total) | 15.87 ± 8.44 | 13.90 ± 8.16 | 13.96 ± 7.20 | 0.258 |
| - Method 4 (Highest frequency) | 3.16 ± 1.15 | 3.20 ± 1.02 | 3.41 ± 0.80 | 0.290 |
| - Method 5 (Highest severity) | 3.19 ± 1.18 | 3.19 ± 1.03 | 3.31 ± 0.87 | 0.721 |

The comparison of day 0 with day 56 showed a significant improvement in each category of SODA scores. Also, the comparison of day 28 and day 56 showed positive changes in each category. The exhaustive numbers for each SODA score were available in Table 3. Furthermore, the detailed analysis of the SODA scores on non-pain symptoms divided according to item 7 to item 13 was available in Table 4. Figures 2, 3, and 4 visually demonstrated changes in SODA scores visually based on pain intensity, nonpain symptoms, and satisfaction, respectively. In all, each summary statistics was displayed by their mean and 95% confidence intervals.

**Table 3.** Comparative Changes in SODA Scores among the C+O, C only, and O only groups.

| SODA Score | Day 0<br>(95% CI) | Day 28<br>(95% CI) | Day 56<br>(95% CI) | p-value | Day 28 – Day 0<br>(95% CI) | p-value | Day 56 – Day 28<br>(95% CI) | p-value | Day 56 – Day 0<br>(95% CI) | p-value |
| --- | --- | --- | --- | --- | --- | --- | --- | --- | --- | --- |
| Pain intensity (2-47) |  |  |  |  |  |  |  |  |  |  |
| ○ C+O | 25.80<br>(24.15, 27.45) | 20.39<br>(18.63, 22.14) | 17.57<br>(15.75, 19.39) | <0.001 | -5.41<br>(-7.37, -3.45) | <0.001 | -2.82<br>(-4.88, -0.76) | 0.007 | -8.23<br>(-10.25, -6.21) | <0.001 |
| ○ C only | 25.14<br>(23.49, 26.8) | 18.92<br>(17.15, 20.69) | 15.55<br>(13.69, 17.42) | <0.001 | -6.22<br>(-8.20, -4.25) | <0.001 | -3.37<br>(-5.48, -1.26) | 0.002 | -9.59<br>(-11.65, -7.53) | <0.001 |
| ○ O only | 26.26<br>(24.6, 27.93) | 19.28<br>(17.52, 21.04) | 15.64<br>(13.76, 17.52) | <0.001 | -6.98<br>(-8.95, -5.02) | <0.001 | -3.64<br>(-5.75, -1.52) | 0.001 | -10.62<br>(-12.70, -8.54) | <0.001 |
| ○ p-value | 0.642 | 0.484 | 0.228 |  | 0.491 |  | 0.914 |  | 0.259 |  |
| Non-pain symptoms (7-35) |  |  |  |  |  |  |  |  |  |  |
| ○ C+O | 16.14<br>(15.40, 16.89) | 13.74<br>(12.95, 14.53) | 11.50<br>(10.68, 12.32) | <0.001 | -2.41<br>(-3.29, -1.52) | <0.001 | -2.24<br>(-3.17, -1.31) | <0.001 | -4.65<br>(-5.56, -3.74) | <0.001 |
| ○ C only | 15.58<br>(14.84, 16.32) | 13.13<br>(12.34, 13.93) | 10.81<br>(9.98, 11.64) | <0.001 | -2.45<br>(-3.34, -1.56) | <0.001 | -2.32<br>(-3.27, -1.38) | <0.001 | -4.77<br>(-5.69, -3.85) | <0.001 |
| ○ O only | 15.74<br>(14.99, 16.48) | 13.18<br>(12.39, 13.98) | 11.15<br>(10.29, 12) | <0.001 | -2.55<br>(-3.44, -1.66) | <0.001 | -2.03<br>(-3, -1.07) | <0.001 | -4.59<br>(-5.53, -3.64) | <0.001 |
| ○ p-value | 0.553 | 0.502 | 0.514 |  | 0.964 |  | 0.791 |  | 0.462 |  |
| Satisfaction (2-23) |  |  |  |  |  |  |  |  |  |  |
| ○ C+O | 11.78<br>(11.15, 12.41) | 12.22<br>(11.54, 12.90) | 12.69<br>(11.98, 13.39) | 0.123 | 0.44<br>(-0.41, 1.28) | 0.309 | 0.47<br>(-0.42, 1.35) | 0.304 | 0.90<br>(0.04, 1.77) | 0.041 |
| ○ C only | 11.83<br>(11.20, 12.47) | 12.70<br>(12.01, 13.38) | 12.88<br>(12.16, 13.60) | 0.038 | 0.86<br>(0.01, 1.72) | 0.048 | 0.19<br>(-0.72, 1.09) | 0.689 | 1.05<br>(0.17, 1.93) | 0.020 |
| ○ O only | 11.68<br>(11.04, 12.31) | 12.41<br>(11.73, 13.08) | 12.74<br>(12.01, 13.47) | 0.049 | 0.73<br>(-0.11, 1.57) | 0.090 | 0.34<br>(-0.57, 1.25) | 0.468 | 1.07<br>(0.18, 1.96) | 0.019 |
| ○ p-value | 0.941 | 0.622 | 0.927 |  | 0.680 |  | 0.921 |  | 0.959 |  |

C, Curcumin; O, Omeprazole

**Table 4.** Comparative Changes in SODA Non-pain Scores among the C+O, C only, and O only groups.

| SODA Score | Day 0<br>(95% CI) | Day 28<br>(95% CI) | Day 56<br>(95% CI) | p-value | Day 28 – Day 0<br>(95% CI) | p-value | Day 56 – Day 28<br>(95% CI) | p-value | Day 56 – Day 0<br>(95% CI) | p-value |
| --- | --- | --- | --- | --- | --- | --- | --- | --- | --- | --- |
| Non-pain symptoms (7-35) |  |  |  |  |  |  |  |  |  |  |
| • Burping/belching |  |  |  |  |  |  |  |  |  |  |
| ○ C+O | 2.10<br>(1.91, 2.30) | 1.79<br>(1.58, 2) | 1.51<br>(1.3, 1.73) | <0.001 | -0.31<br>(-0.55, -0.07) | 0.011 | -0.28<br>(-0.53, -0.02) | 0.032 | -0.59<br>(-0.83, -0.34) | <0.001 |
| ○ C only | 2.16<br>(1.96, 2.36) | 1.81<br>(1.60, 2.02) | 1.57<br>(1.35, 1.79) | <0.001 | -0.35<br>(-0.59, -0.11) | 0.004 | -0.24<br>(-0.49, 0.02) | 0.068 | -0.59<br>(-0.84, -0.34) | <0.001 |
| ○ O only | 2.06<br>(1.86, 2.26) | 1.80<br>(1.59, 2.01) | 1.47<br>(1.24, 1.69) | <0.001 | -0.26<br>(-0.50, -0.02) | 0.036 | -0.33<br>(-0.59, -0.08) | 0.011 | -0.59<br>(-0.84, -0.34) | <0.001 |
| • Heartburn |  |  |  |  |  |  |  |  |  |  |
| ○ C+O | 2.49<br>(2.27, 2.72) | 1.82<br>(1.58, 2.06) | 1.29<br>(1.04, 1.54) | <0.001 | -0.67<br>(-0.96, -0.39) | <0.001 | -0.53<br>(-0.83, -0.23) | 0.001 | -1.20<br>(-1.50, -0.91) | <0.001 |
| ○ C only | 2.10<br>(1.88, 2.33) | 1.54<br>(1.30, 1.79) | 1.30<br>(1.04, 1.55) | <0.001 | -0.56<br>(-0.85, -0.27) | <0.001 | -0.25<br>(-0.55, 0.06) | 0.118 | -0.80<br>(-1.10, -0.50) | <0.001 |
| ○ O only | 2.19<br>(1.96, 2.42) | 1.43<br>(1.19, 1.68) | 1.36<br>(1.10, 1.62) | <0.001 | -0.76<br>(-1.04, -0.47) | <0.001 | -0.08<br>(-0.39, 0.23) | 0.620 | -0.83<br>(-1.14, -0.53) | <0.001 |
| • Bloating |  |  |  |  |  |  |  |  |  |  |
| ○ C+O | 2.72<br>(2.48, 2.97) | 1.91<br>(1.66, 2.17) | 1.79<br>(1.52, 2.06) | <0.001 | -0.81<br>(-1.11, -0.51) | <0.001 | -0.13<br>(-0.44, 0.19) | 0.429 | -0.94<br>(-1.25, -0.63) | <0.001 |
| ○ C only | 2.72<br>(2.48, 2.97) | 2.01<br>(1.75, 2.27) | 1.48<br>(1.21, 1.75) | <0.001 | -0.72<br>(-1.02, -0.42) | <0.001 | -0.53<br>(-0.85, -0.21) | 0.001 | -1.25<br>(-1.56, -0.93) | <0.001 |
| ○ O only | 2.85<br>(2.61, 3.1) | 2.12<br>(1.86, 2.38) | 1.72<br>(1.44, 2) | <0.001 | -0.73<br>(-1.04, -0.43) | <0.001 | -0.40<br>(-0.72, -0.08) | 0.015 | -1.13<br>(-1.45, -0.82) | <0.001 |
| • Passing gas |  |  |  |  |  |  |  |  |  |  |
| ○ C+O | 2.23<br>(2.04, 2.43) | 1.91<br>(1.70, 2.12) | 1.57<br>(1.35, 1.79) | <0.001 | -0.32<br>(-0.58, -0.06) | 0.016 | -0.34<br>(-0.61, -0.07) | 0.014 | -0.66<br>(-0.93, -0.39) | <0.001 |
| ○ C only | 2.01<br>(1.82, 2.21) | 1.67<br>(1.46, 1.88) | 1.35<br>(1.13, 1.58) | <0.001 | -0.35<br>(-0.61, -0.09) | 0.009 | -0.32<br>(-0.59, -0.04) | 0.026 | -0.66<br>(-0.93, -0.39) | <0.001 |
| ○ O only | 2.01<br>(1.82, 2.21) | 1.91<br>(1.70, 2.12) | 1.48<br>(1.26, 1.71) | <0.001 | -0.11<br>(-0.37, 0.15) | 0.413 | -0.42<br>(-0.70, -0.14) | 0.003 | -0.53<br>(-0.80, -0.26) | <0.001 |
| • Sour Taste |  |  |  |  |  |  |  |  |  |  |
| ○ C+O | 1.75<br>(1.58, 1.93) | 1.40<br>(1.21, 1.58) | 1.23<br>(1.04, 1.42) | <0.001 | -0.36<br>(-0.58, -0.13) | 0.002 | -0.17<br>(-0.40, 0.07) | 0.165 | -0.52<br>(-0.75, -0.29) | <0.001 |
| ○ C only | 1.70<br>(1.52, 1.87) | 1.27<br>(1.08, 1.46) | 1.05<br>(0.85, 1.24) | <0.001 | -0.43<br>(-0.65, -0.20) | <0.001 | -0.22<br>(-0.46, 0.02) | 0.068 | -0.65<br>(-0.88, -0.42) | <0.001 |
| ○ O only | 1.71<br>(1.53, 1.88) | 1.48<br>(1.29, 1.67) | 1.28<br>(1.08, 1.49) | 0.002 | -0.23<br>(-0.45, 0) | 0.049 | -0.20<br>(-0.44, 0.04) | 0.111 | -0.42<br>(-0.66, -0.19) | <0.001 |
| • Nausea |  |  |  |  |  |  |  |  |  |  |
| ○ C+O | 1.77<br>(1.61, 1.92) | 1.36<br>(1.19, 1.53) | 1.14<br>(0.96, 1.31) | <0.001 | -0.41<br>(-0.61, -0.20) | <0.001 | -0.22<br>(-0.44, -0.01) | 0.044 | -0.63<br>(-0.84, -0.42) | <0.001 |
| ○ C only | 1.67<br>(1.51, 1.82) | 1.22<br>(1.05, 1.39) | 1.09<br>(0.91, 1.26) | <0.001 | -0.45<br>(-0.66, -0.24) | <0.001 | -0.13<br>(-0.35, 0.09) | 0.246 | -0.58<br>(-0.80, -0.37) | <0.001 |
| ○ O only | 1.60<br>(1.44, 1.76) | 1.21<br>(1.04, 1.38) | 1.13<br>(0.94, 1.31) | <0.001 | -0.39<br>(-0.60, -0.18) | <0.001 | -0.09<br>(-0.31, 0.14) | 0.453 | -0.48<br>(-0.70, -0.26) | <0.001 |
| • Bad Breath |  |  |  |  |  |  |  |  |  |  |
| ○ C+O | 1.65<br>(1.50, 1.81) | 1.16<br>(0.99, 1.32) | 1.08<br>(0.90, 1.25) | <0.001 | -0.50<br>(-0.70, -0.30) | <0.001 | -0.08<br>(-0.29, 0.13) | 0.455 | -0.58<br>(-0.78, -0.37) | <0.001 |
| ○ C only | 1.54<br>(1.38, 1.69) | 1.21<br>(1.05, 1.38) | 1.11<br>(0.94, 1.29) | <0.001 | -0.33<br>(-0.53, -0.12) | 0.001 | -0.10<br>(-0.31, 0.12) | 0.368 | -0.42<br>(-0.63, -0.22) | <0.001 |
| ○ O only | 1.53<br>(1.37, 1.68) | 1.27<br>(1.11, 1.44) | 1.1<br>(0.92, 1.28) | <0.001 | -0.26<br>(-0.46, -0.06) | 0.012 | -0.17<br>(-0.39, 0.04) | 0.117 | -0.43<br>(-0.64, -0.22) | <0.001 |

C, Curcumin; O, Omeprazole

**Figure 2.**
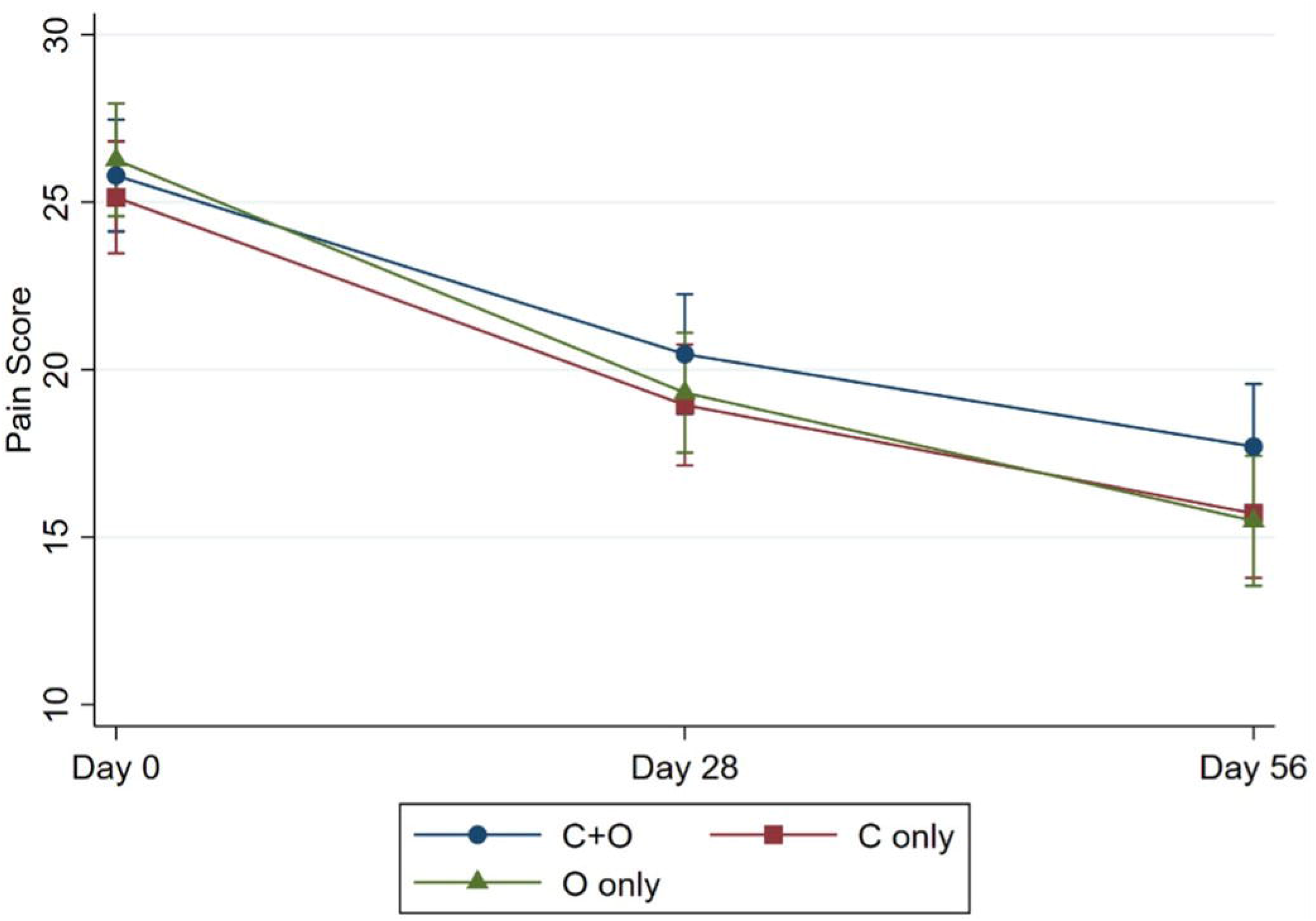
Comparative Changes in SODA Pain Scores.

**Figure 3.**
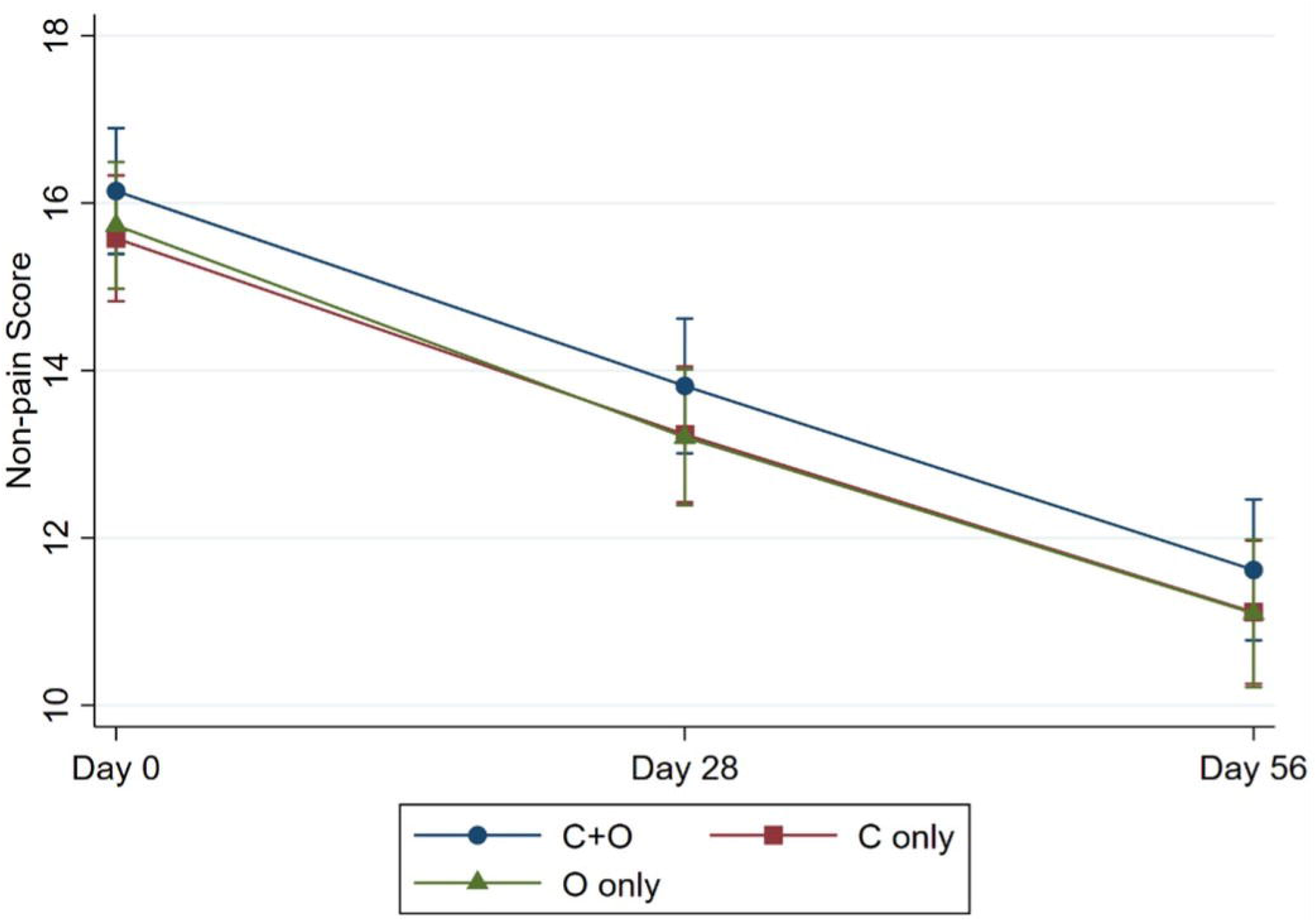
Comparative Changes in SODA Nonpain Scores.

**Figure 4.**
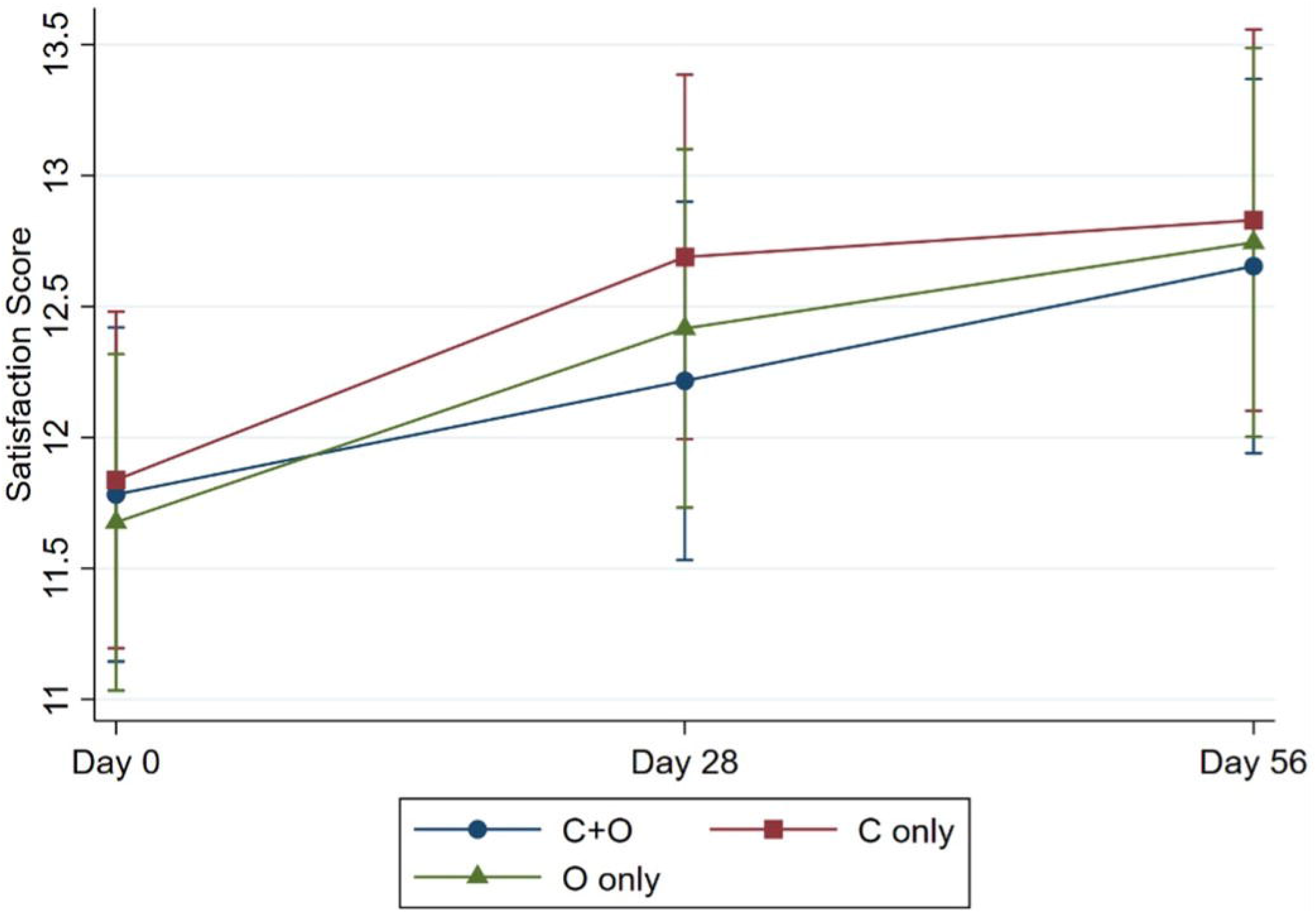
Comparative Changes in SODA Satisfaction Scores.

Analysis of SODA scores by a GEE did not show significant differences between the C+O group versus the C group, the C+O group versus the O group, and the C group versus the O group (Table 5).

**Table 5.** Pairwise Comparison of SODA Scores.

| SODA Scores | C+O vs C only<br>(95% CI) | p-value | C+O vs O only<br>(95% CI) | p-value | C only vs O only<br>(95% CI) | p-value |
| --- | --- | --- | --- | --- | --- | --- |
| Pain intensity (2-47) | -1.23 (-3.09, 0.64) | 0.197 | -0.70 (-2.57, -1.17) | 0.461 | 0.52 (-1.35, 2.40) | 0.584 |
| Non-pain symptoms (7-35) | -0.54 (-1.37, 0.28) | 0.196 | -0.42 (-1.25, 0.40) | 0.316 | 0.12 (-0.71, 0.95) | 0.776 |
| Satisfaction (2-23) | 0.22 (-0.41, 0.86) | 0.486 | 0.03 (-0.60, 0.66) | 0.929 | -0.20 (-0.83, 0.44) | 0.546 |

C, Curcumin; O, Omeprazole

### Adverse events

Adverse events included anxiety, diarrhea, drowsiness, flatulence, headache, and vomiting. In the group C+O, these occurred in two patients (2.90%), including one diarrhea (1.45%), one drowsiness (1.45%), one headache (1.45%), and one vomiting (1.45%). In Group C only, these occurred in three patients (4.35%), including one incident of anxiety (1.45%), one diarrhea (1.45%), one flatulence (1.45%), and one vomiting (1.45%). No serious adverse events took place.

## Discussion

Curcumin is a low molecular weight hydrophobic polyphenol extracted from turmeric. Curcumin possesses a wide spectrum of biological properties, including anti-inflammatory, antioxidant, antiproliferative, and antimicrobial properties. Numerous clinical trials have established the pharmacological properties of curcumin. Mechanisms of symptom generation in patients with functional digestive disorders are poorly understood, due to the absence of a mucosal injury that could explain their distressing symptoms (10). Transient receptor potential vanilloid type 1 (TRPV1) receptors have been shown to play a critical role in the detection and transmission of somatic and visceral nociceptive neural signals(11), and have been implicated in the induction of symptoms in these diseases. TRPV1 is a multimodal sensory transducer that can be activated by a variety of harmful stimuli including heat, low pH, endogenous lipid derivatives such as anandamide, and exogenous vanilloid-containing substances such as capsaicin (12). In particular, curcumin shares the same vanilloid ring moiety as capsaicin, making TRPV1 a likely target, and it has been shown in animals that curcumin inhibits capsaicin-induced TRPV1 activation competitively (13). Increased TRPV1 signaling has been suggested to contribute to visceral hypersensitivity in functional gastrointestinal diseases, including esophageal hypersensitivity(13). Additionally, TRPV1 receptors are highly expressed throughout the gastrointestinal tract and enteric nervous system, and there is evidence that curcumin can inhibit gastrointestinal nociception and reverse intestinal hypersensitivity through peripheral terminals. With this mechanism of action in mind, it cannot be ruled out that this molecule may be beneficial in the treatment of patients with functional dyspepsia and irritable bowel syndrome, which are disorders that remain clinically challenging in the presence of currently available medications and whose patients may benefit from curcumin’s pharmacological properties on TRPV1 as a novel pain modulator. Curcumin has been clinically studied in patients with inflammatory bowel disease, irritable bowel syndrome, ulcers, *Helicobacter pylori* infections, and even pancreatitis. Curcumin is effective in the treatment of gastric ulcers, erosions, and dyspepsia (14, 15), with ulcers and erosions reduced or even eradicated after administration of curcumin (3,000 mg/day) administration for up to 12 weeks, while abdominal pain and discomfort were significantly reduced. This explains why in this study we compared curcumin with a PPI as a treatment for functional dyspepsia.

Similarly to the findings of the current study, curcumin is safe in numerous human studies, with only minor toxicity associated with this polyphenol(16). Velayudhan et al. also documented the traditional use of curcumin and noted that even a single oral dose of up to 8000 mg was not detected in the serum(17). Therefore, curcumin is increasingly being viewed as a biomolecule capable of being administered for an extended period without causing adverse effects(18). After 72 hours, safety was assessed in a dose increase study involving 34 healthy volunteers who received curcumin doses ranging from 500 to 12,000 mg. Only seven subjects reported mild disturbances, including headache, skin rash, diarrhea, and yellow stool(19). Another study, which lasted 1-4 months, found that increasing the dose of curcumin from 0.45 to 3.6 g/d resulted in rare cases of nausea and diarrhea, as well as increased alkaline phosphatase and lactate dehydrogenase (20). Some patients treated with doses as high as 8 g/d for two weeks complained of abdominal pain and bulky size (21). The findings of the current study confirmed the safety of curcumin compared to PPIs when used to treat functional dyspepsia.

Recently published Cochrane Database of Systematic Reviews demonstrated that PPI was more effective than placebo in the treatment of functional dyspepsia, regardless of the dose or duration of treatment (6, 22). The presence of reflux symptoms or different subtypes of functional dyspepsia had no effect on the effect of PPI over placebo. PPIs may be slightly more effective in alleviating general symptoms of dyspepsia than H2RA and prokinetics. However, several previous studies have demonstrated adverse events associated with long-term PPI use (23, 24). Therefore, trials are required that examine the longer-term benefits and harms (at least six to twelve months) benefits and harms of PPI in functional dyspepsia. To expand the treatment options for patients with functional dyspepsia, we decided to compare PPI and curcumin in this study. The findings of the current study indicate that there are no significant adverse events associated with the short-term use of PPI and curcumin.

To our knowledge, this is the first study to demonstrate the efficacy of curcumin in functional dyspepsia. Curcumin is effective in all subtypes of functional dyspepsia. Curcumin and omeprazole are both effective for functional dyspepsia and do not appear to have a synergistic effect. Although the most recent Thailand dyspepsia guidelines (2018) (28) recommended that patients with undiagnosed dyspepsia who do not have alarm symptoms receive an empirical trial of PPI for 4-8 weeks as first-line therapy. Prokinetic agents may be used in patients with unexplained dyspepsia who do not improve after empirical PPI therapy. Furthermore, prokinetic agents, tricyclic antidepressants, and cytoprotective agents have been shown to improve symptoms in patients with functional dyspepsia after the failure of PPI therapy. Although this guideline did not specifically mention curcumin as a treatment option for functional dyspepsia, the new findings from our study may justify considering curcumin in clinical practice.

As a multicentre randomized controlled trial, this study is highly reliable. The study’s subject is also noteworthy because functional dyspepsia is a prevalent disorder in the general population. The chosen drug is a proton pump inhibitor, widely used, has been approved for over-the-counter use, and is the first-line therapy for patients with functional dyspepsia. The medication chosen as a comparator is curcumin, a popular herbal remedy in the general population. In the context of gastrointestinal disorders, there is no precedent for comparing two drugs in a double-blind randomized controlled trial. The study participants met all criteria and were also diagnosed separately by endoscopy to rule out the presence of symptoms consistent with other diseases. They were also tested for H Pylori and those with infection were isolated.

In addition, the research methods are completely bias-free, as the individuals who administered the drugs, the participants who received the drugs, and the individuals who performed the assessment were unaware of the type of medications taken by the participants. The trial was carried out in hospitals frequented by the participants, namely hospitals in other provinces and Thai traditional medicine hospitals. The assessment used standardized questionnaires, and the individuals who conducted it were also certified for accuracy in the assessment. The number of participants was determined statistically accurately using standardized research methods.

The number of participants and the randomization used to assign them to different groups were conducted in a confidential and unbiased manner. All individuals involved in the drug administration process were unaware of the types of drugs distributed or to whom they were distributed. The statistical tests used in this study were conducted using appropriate materials and according to accepted statistical principles.

Two additional follow-up appointments have been scheduled to address the side effects of the study. Blood tests were performed on participants during the second follow-up to assess liver function. No abnormal symptoms were observed after the administration of either of the drugs.

The study findings indicated that the required number of participants was reached. However, it was discovered that several participants did not provide follow-up information after medication administration, which is the weakness of the study. However, the number of participants who provided this information was sufficient for statistical analysis and the majority of the participants attended the follow-up. Therefore, it can be deduced from the results that even if the number of participants followed after drug administration increased, the study findings would not be significantly different. Another limitation of this study is the absence of long-term follow-up data for all patients after treatment. This is a question that will require further investigation.

The strength of the study is that the findings can be applied to patients with functional dyspepsia who visit their doctor in general clinics or hospitals since the study settings correspond to the daily operations of physicians who already encounter this group of patients. According to the research findings, patients with functional dyspepsia would have additional drug options in addition to proton pump inhibitors alone, with no additional side effects. The study findings can be made completely public. The study was partially funded by government organizations, ensuring that there was no bias in the selection of particular medications. Additionally, this study is the first well-designed randomized controlled trial of curcumin versus PPI in functional dyspepsia. Functional dyspepsia was confirmed by endoscopy and *H. pylori* infection was ruled out. Although we strictly exercised the method to ensure the high integrity of the experiment, this study was subject to limitations, including a small number of patients who were lost to follow-up and a lack of long-term follow-up data.

Future studies in this issue will examine the long-term benefit and harms (at least 6-12 months) benefit and harms of curcumin in functional dyspepsia; the results of the use of curcumin on demand long-term in functional dyspepsia; and the efficacy of curcumin in other functional gastrointestinal disorders.

## Conclusion

Curcumin and omeprazole have comparable efficacy for functional dyspepsia with no obvious synergistic effect.

## Data Availability

All data produced in the present study are available upon reasonable request to the authors.

## Abbreviations

AE: Adverse Events
C: Curcumin
C+O: Curcumin+Omeprazole
EPS: Epigastric Pain Syndrome
O: Omeprazole
PDS: Postprandial Distress Syndrome
PPIs: Proton Pump Inhibitors
SAE: Serious Adverse Events
SF-LDQ: Short-Form Leeds Dyspepsia Questionnaire
SODA: Severity of Dyspepsia Assessment (SODA)

## Author’s Contributions

- Pradermchai Kongkam: Data curation, Writing original draft, Review and Editing
- Wichittra Khongkha: Data curation, Writing original draft, Review and Editing
- Chawin Lopimpisuth: Writing original draft, Review and Editing
- Chisanucha Chumsri: Writing original draft, Review and Editing
- Prach Kosarussawadee: Data curation, Review and Editing
- Phanupong Phutrakool: Data curation, Review and Editing
- Sittichai Khamsai: Data curation, Review and Editing
- Kittisak Sawanyawisuth: Data curation, Review and Editing
- Thanyachai Sura: Data curation, Review and Editing
- Pochamana Phisalprapa: Data curation, Review and Editing
- Thanwa Buamahakul: Data curation, Review and Editing
- Sarawut Siwamogsatham: Data curation, Review and Editing
- Jaenjira Angsusing: Administration, Data curation, Review and Editing
- Pratchayanan Poonniam: Administration, Data curation, Review and Editing
- Kulthanit Wanaratna: Supervision, Data curation, Review and Editing
- Monthaka Teerachaisakul: Supervision, Funding, Data curation, Review and Editing
- Krit Pongpirul: Study Design, Data curation, Formal Analysis, Writing original draft, Review and Editing

